# Generalizable and Automated Classification of TNM Stage from Pathology Reports with External Validation

**DOI:** 10.1101/2023.06.26.23291912

**Authors:** Jenna Kefeli, Nicholas Tatonetti

## Abstract

Cancer staging is an essential clinical attribute informing patient prognosis and clinical trial eligibility. However, it is not routinely recorded in structured electronic health records. Here, we present a generalizable method for the automated classification of TNM stage directly from pathology report text. We train a BERT-based model using publicly available pathology reports across approximately 7,000 patients and 23 cancer types. We explore the use of different model types, with differing input sizes, parameters, and model architectures. Our final model goes beyond term-extraction, inferring TNM stage from context when it is not included in the report text explicitly. As external validation, we test our model on almost 8,000 pathology reports from Columbia University Medical Center, finding that our trained model achieved an AU-ROC of 0.815-0.942. This suggests that our model can be applied broadly to other institutions without additional institution-specific fine-tuning.

## Introduction

Cancer stage, an important diagnostic and prognostic clinical attribute, is frequently used to identify patients for clinical trial recruitment and research cohort construction. While not routinely captured in the electronic health record, stage information can be found in patient pathology reports. Tumor registries, tasked with manually identifying stage from clinical notes and pathology reports, can take up to 6 months from diagnosis to extraction, at which point the opportunity for clinical trials or other treatments may have passed [1-2]. A shortage of cancer registry specialists suggests that this lead time may become even longer [3]. In this study, we present a transformer-based method for the automated classification of TNM stage from pathology report text across 23 cancer types. Transformer-based methods have been applied to other clinical text [4], but have not been widely applied to pathology reports. We demonstrate that our model is generalizable to an independent institution, suggesting other institutions can use our method in an “off-the-shelf” capacity.

Extraction of cancer stage has been an ongoing effort. Previous studies have focused on single cancer types [5-6], used smaller-sized training [5-6] or testing [5-7] data sets (<1,000 reports), and have relied on single-institution data without external validation [6-7]. Some studies required additional data beyond pathology report text as model input [5, 8]. For methods, two studies employed regular expression and customized rule-based approaches [6-7], one utilized traditional machine learning methods [5], and another used a hybrid transformer-embedding and deep learning model [8]. The majority of prior work classified patients into broad TNM categories that do not cover the entire spectrum of clinical values [5-6, 8], whereas in this study we classify reports into more granular, clinically relevant TNM categorizations. This is a more difficult classification task, as each TNM category has a greater number of possible outcomes. Finally, none of the previous research studies have made their models publicly available, whereas we are releasing our trained TNM models to be directly utilized by other institutions.

Here, we utilize recent advances in natural language processing to classify cancer stage directly from pathology report text [9]. We specifically use a new variant of BERT [10-11], which has a larger input capacity than previous versions, and show that our model performs better than standard BERT models. Indeed, increased input capacity is important for subsequent deployment of trained models, as real-world documents are typically longer than the limited input capacity of standard models. To our knowledge, this is the first application of high-input capacity large language models (LLMs) to pathology report text for classification of any prediction target.

## Results

Our overall approach consisted of (1) training a model using publicly available pathology reports and then (2) applying our trained model to a set of independent reports for validation of generalizability (Figure 1A). Cancer stage is comprised of three elements: tumor size (T), regional lymph node involvement (N), and distant metastasis (M). Our prediction task consisted of classifying reports into TNM staging categories, with a separate model trained for each variable.

**Figure 1.**
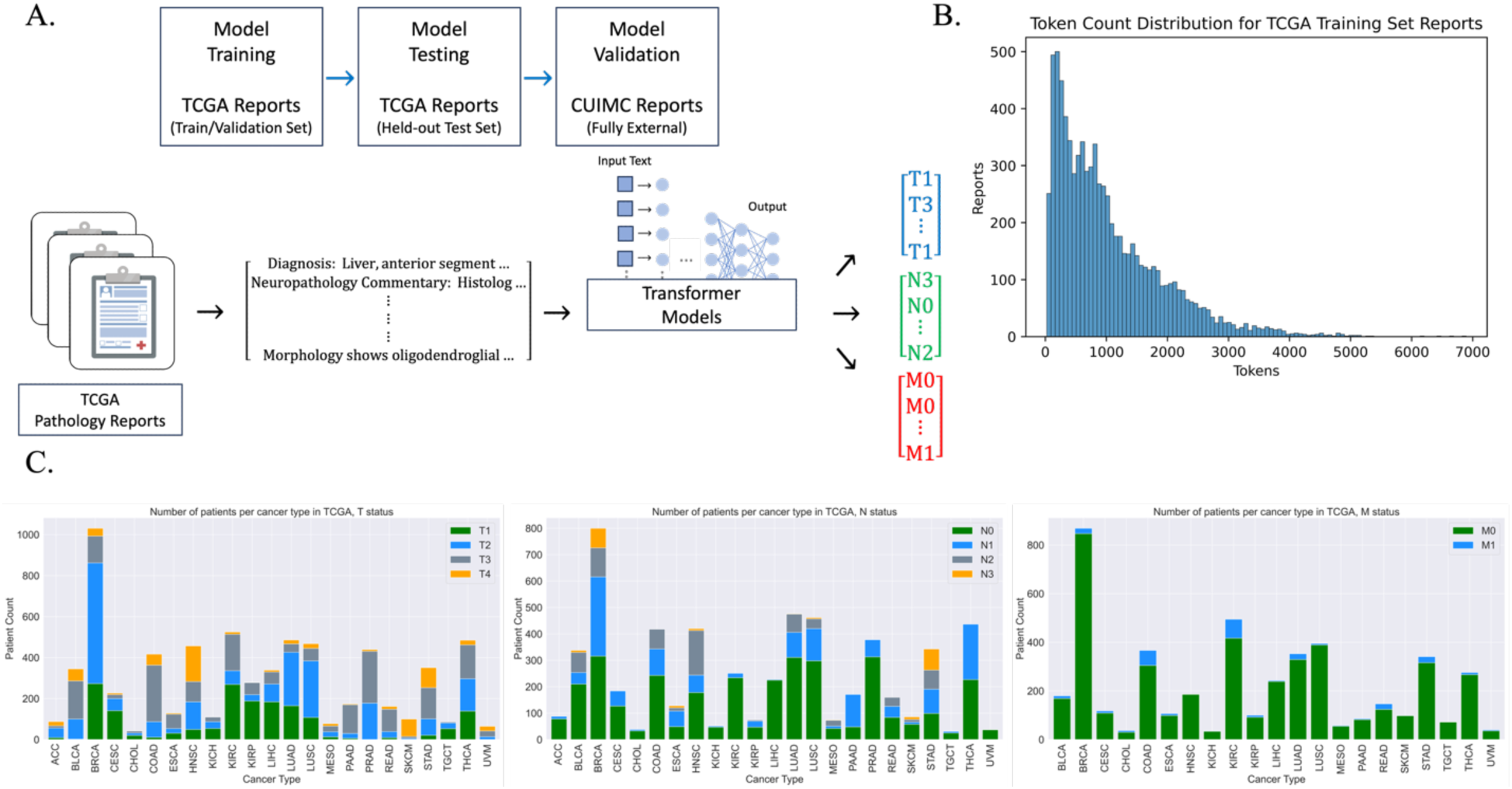
**(A)** Depiction of overall method. Top: Dataset separation into training/validation and held-out test sets (TCGA), as well as external validation (CUIMC). Bottom: Example TCGA pathology reports, inputted into separate transformer models to for TNM stage prediction. **(B)** Token distribution for TCGA training set reports. The ClinicalBERT (CB) tokenizer was used to tokenize reports into pre-defined CB vocabulary. (**C)** Per-class distribution of TCGA pathology reports with TNM staging annotation. The distribution of TNM values varied substantially between cancer types. X-axis labeled as TCGA cancer-type abbreviations.

First, we selected 9,523 pathology reports from The Cancer Genome Atlas (TCGA) [9, 12]. The availability of TNM annotation in the TCGA metadata varied: 6,887 reports were documented with known tumor size (T), 5,678 reports with known regional lymph node involvement (N), and 4,608 reports with known metastasis (M). All TNM values were based on pathologist review of relevant histology slides. Patients with known T and N values spanned 23 different cancer types, while patients with known M status corresponded to 21 cancer types. Due to the large number of contributing pathologists and institutions represented in the TCGA dataset (Figure S1), we observed that report structure, composition, and terminology varied greatly, even amongst single cancer type subsets.

The complexity and size of the dataset suggested that a large language model (LLM) would be appropriate, and that such a model should be generalizable once trained. To this end, we tested two pre-trained LLMs for our classification task [11, 13] (Methods). Both models were pre-trained on a large set of publicly available clinical notes [14]. The first, ClinicalBERT (CB) [13], has been widely used in the field of clinical natural language processing, but is limited in both training and application by its maximum input capacity (512 tokens per document). Indeed, we found that over 66% of TCGA reports are greater than 512 tokens in length (Figure 1B, Table S1). The second model, Clinical-BigBird (CBB) [11], recently released, has a vastly increased document input capability (4,096 tokens per document) with proportionally fewer model parameters. We implemented both models to compare performance on our TNM classification tasks.

Classification was divided into individual tasks due to differences in patient training set size based on TNM status. Each classification target was assigned different integer-value ranges based on standard clinical use: T-values were in [1, 2, 3, 4], N-values were in [0, 1, 2, 3], and M-values were binary, [0, 1]. We denote these ranges as T14, N03, and M01 (Figure 1C). We divided TCGA patients into training, validation, and held-out test sets. We selected the best-performing model for each target based on validation set optimization. The model-type that performed best across all three targets was CBB. We varied input size, finding that CBB models parameterized with larger input sizes generally performed better – CBB with 2,048 input tokens performed best for T14 and N03 targets, and CBB with 1,024 input tokens for the M01 target (Table S2). Validation set performance ranged from 0.823-0.959 AU-ROC. We then evaluated best-model performance on TCGA held-out test sets, with AU-ROC ranging from 0.750– 0.945 (Figure S2-3). The TCGA test sets were completely independent of the training dataset for each target.

We further evaluated our best-performing models on an independent set of pathology reports. We selected all pathology reports from Columbia University Irving Medical Center (CUIMC) from 2010-2019, and matched with tumor registry TNM annotation based on report date and diagnosis (see Methods). As in the TCGA dataset, there was uneven coverage of TNM annotation across patients: 7,792 patients corresponded to known T status, 6,140 patients corresponded to known N status, and 2,245 patients corresponded to known M status (Table S3). Patients with T status spanned 42 primary cancer sites; patients with N status spanned 41 primary cancer sites, and patients with M status spanned 40 primary cancer sites (Figure S4).

The final models were not fine-tuned on CUIMC reports, but rather applied directly in an “off the shelf” capacity. We found that our CBB models performed well, with AU-ROC ranging from 0.815-0.942 (Figure 2A-D). For the multi-class targets T14 and N03, we found that per-class performance was consistently high (Figure 2A-B). To ascertain whether the use of the CBB model-type (with its increased complexity and much larger input size) made a difference in application to CUIMC data, we compared the best-performing CB model (as determined on the TCGA validation set) to the best-performing CBB model for the N03 task. We found that the best CB model performed at AU-ROC of 0.779, whereas the best CBB model produced AU-ROC 0.912 on CUIMC data (Table S4).

**Figure 2.**
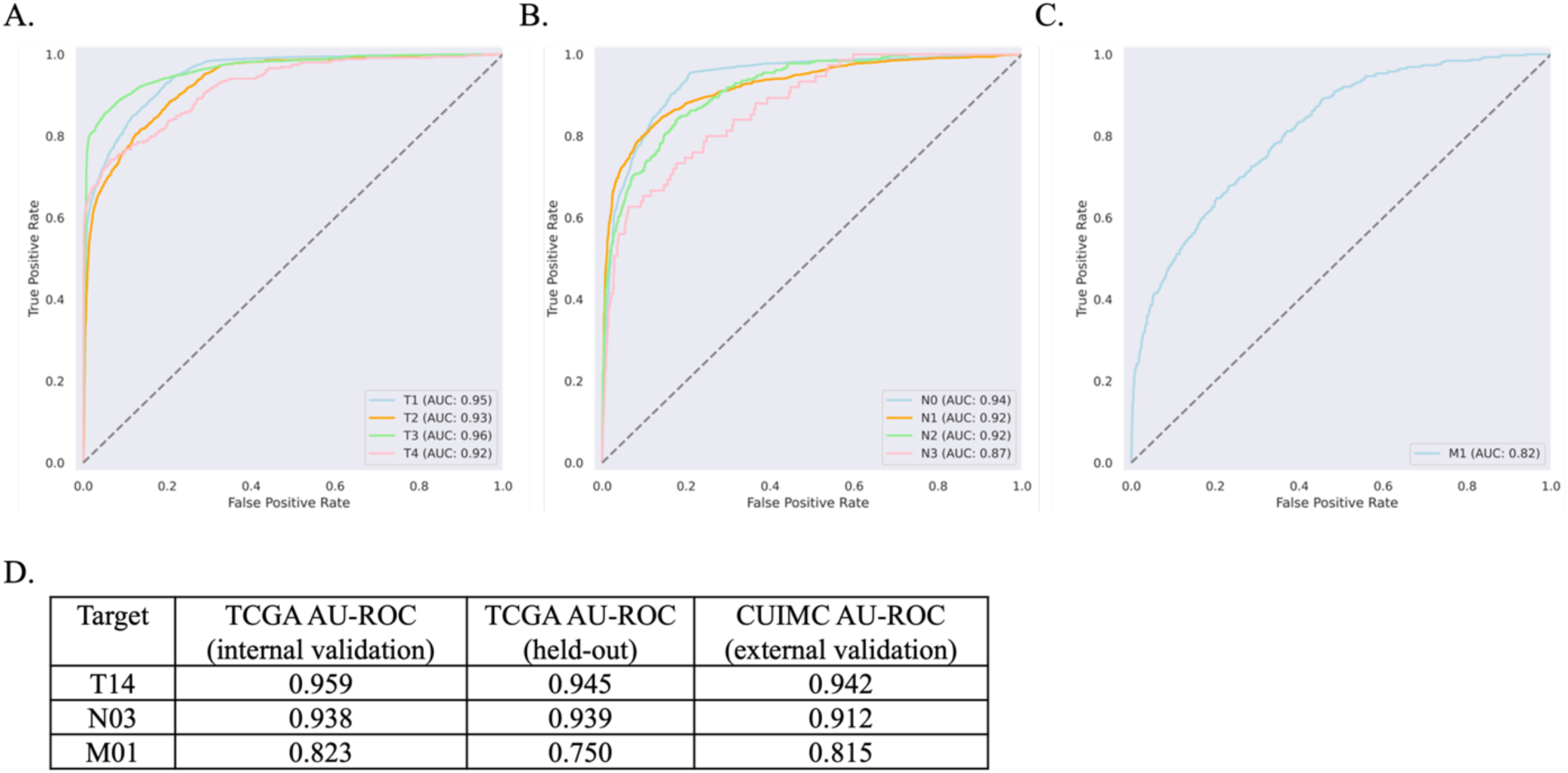
Performance of final models. **(A-C)** External validation using CUIMC data. Per-Class ROC Curves for each model: Tumor size (T14), Reginal lymph node involvement (N03), and Distant metastasis (M01). (**D)** Best-performing models applied to TCGA held-out and CUIMC pathology reports. Models were selected based on TCGA internal validation set performance.

Finally, we tested whether including protected health information (PHI), such as name, date of birth, MRN, and gender, would have any impact on model performance for T14. We found that the best-performing model had a very slight improvement in performance (difference in AU-ROC of 0.0001) when PHI was removed (Table S5). Given the small difference in performance, we do not consider PHI exclusion to be a necessary requirement for the use or application of our models.

## Discussion

Automating the classification of cancer stage from pathology reports would result in shorter turn-around time for clinical trial patient selection and research cohort construction. This would allow more patients to be routinely surveyed for inclusion in clinical trials and research studies. Automated stage annotation may be utilized in facilitating automated pathology report review for clinical use and prognostication, and in developing multi-modal models that combine both image and text, particularly as medical centers are increasingly digitizing pathology slides [15].

In this study, we applied a recently released transformer model, with higher input capacity than the traditional BERT model, to achieve consistently high predictive performance across multiple independent datasets. Importantly, we have made our models publicly available. Unlike other studies in clinical NLP, our models were trained on fully de-identified pathology reports, and therefore do not necessitate specialized approaches, such as differential privacy or federated learning, for their dissemination. As we have demonstrated, the TNM models are consistently performant across institutions. We found that our TCGA-trained models performed as well on the CUIMC report set as on the TCGA held-out test set (Figure 2D), suggesting that they are indeed generalizable. Many CUIMC reports did not contain explicit staging terms (15.3% of T14, 20.3% of N03, and 94.7% of M01 reports), suggesting that the models exhibit increased performance by going beyond direct extraction, inferring stage classification from context. Future work should include additional testing at external institutions to further validate the generalizability of our TNM models.

Although promising, our method has a number of notable limitations. Firstly, we abstracted the values of T, N, and M, converting for example “T1a” to “T1.” Re-training our models on a larger set of pathology reports would allow for a more detailed prediction target. Second, we were limited by our computational memory allotment, restricting the number of tokens per report to 2,048. If computational capacity were increased, one may extend to 4,096 tokens in order to capture longer reports at full-length. Third, we developed our models based on current AJCC [16] definitions of stage; our model would need to be re-trained if AJCC definitions were substantially updated.

In addition, the M01 model did not perform as well as the T14 and N03 models overall. This is likely due to a number of factors inherent to the TCGA training data: (1) M01 was a particularly imbalanced dataset, with only 6.7% of reports having M1 annotation (Figure 1C) due to TCGA design preference for non-metastatic cases, (2) many reports did not contain M0 or M1 explicitly, as compared to the other targets (95.1% of reports did not contain explicit M01, as compared to 66.5% for N03 and 35.8% for T14), and (3) TCGA annotations for M01 were at times inconsistent with report text (see Methods). The M01 model is limited by the quality of the input data on which it is trained. In future work, the M01 model may be improved by training on a dataset with a great number of M1-annotated pathology reports and more consistent ground truth annotation.

## Methods

### TCGA Pathology Report Dataset Construction with TNM Annotation

Pathology reports and associated TNM clinical metadata were downloaded from the TCGA Genomic Data Commons (GDC) data portal [17]. Reports were initially stored in PDF format; in previous work, we converted the TCGA pathology report corpus to machine-readable plain text using OCR, performed extensive curation, and fully characterized the final TCGA report set. The final dataset spanned 9,523 reports, with 1:1 patient:report ratio [9, 18].

TNM staging annotation was contained within the clinical metadata provided by TCGA [17]. The TNM staging attribute used in this study is “pathological stage,” i.e., stage based on pathologist assessment of patient tumor slide(s). TNM values were abstracted to numerical values, without additional letter suffixes -For example, “N1B” was converted to “N1.” Data availability, or TNM coverage, varied. A given report may have had no associated TNM data, full associated TNM data, or some combination of associated TNM values. Due to the difference in coverage, we separated the data by TNM data availability for individual classification tasks. Each target dataset consisted of non-uniform target value distributions, as displayed in Figure 1C, to varying degrees.

Finally, TCGA annotation of M01 was found to be inconsistent. We examined a random sample of 10 pathology reports, with 5 reports annotated as M0 and 5 reports annotated as M1 in the TCGA metadata. We found that 5/5 reports annotated as M0 were labeled consistently with the AJCC definition of M0. However, we found that 2/5 reports annotated as M1 were not labeled consistently with the AJCC definition of M1 (“distant metastasis”), but rather contained characteristics similar to the reports labeled M0. From this, we observe that the ground truth labels for the M01 target may not be uniformly accurate, as they we found to be at times inconsistent with the AJCC definitions of distant metastasis and inconsistently applied among reports.

### Comparison of Clinically Pre-Trained BERT-Based Models

For each target, we performed fine-tuning experiments using two model-types, ClinicalBERT [13] and Clinical-BigBird [11]. Both models had been pre-trained on a set of clinical notes (MIMIC III [14]). ClinicalBERT (CB) has consistently performed at a high level across a variety of clinical natural language processing tasks [19-21]. Model CB contains 108.3M parameters and is based on the classic BERT architecture [10]. CB is, however, very limited by a maximum input document length of 512 tokens. As a result, reports longer than 512 tokens are truncated during training, and text beyond 512 tokens is not used for model learning. In addition, when applying the model to an external dataset, reports must again be truncated to 512 tokens, so that any information contained within text beyond 512 tokens is not applied toward model prediction. As many real-world reports are longer than 512 tokens, this is a serious limitation.

A more recent model, Clinical-BigBird (CBB), has 128.1M parameters and adopts the computationally-optimized *BigBird* architecture [22]. Bigbird is based on the BERT architecture, but differs in the specification of the attention mechanism. Briefly, a sparse attention mechanism allows for longer inputs to be computationally tractable, providing linear run-time with number of input tokens (compared to the quadratic run-time of BERT) and better performance on benchmark tasks [22]. As a result, model CBB has a vastly increased document length capacity (4,096 tokens), which is allows the use of entire-length reports in both training and application. For example, in the TCGA pathology report dataset, over 66% of reports in the TCGA dataset contain > 512 tokens (Table S1), while 12.9% have report length > 2,048 tokens, and only 0.7% have reports > 4,096 tokens.

### Multi-Class Classification Tasks utilizing the TCGA Pathology Report Dataset

We separated reports into reports with M01 annotation, reports with N03 annotation, and reports with T14 annotation. M01 annotation had the least coverage in the TCGA dataset overall. Each report set was divided into training (70%), validation (15%), and held-out test (15%) sets. As each patient corresponded to a single report, no patient spanned more than one train/validation/test (TVT) subset. In addition, when separating the reports into TVT subsets, we balanced on TNM value composition so that the same balance of values was consistent across TVT subsets. This allowed for fair comparison of performance across TVT subsets, with no TVT subset having a greater imbalance than the dataset overall.

Independent models were trained and hyperparameter-optimized for each of M01, N03, and T14 classification targets separately, as specified below. We evaluated model performance based on macro AU-ROC and per-class AU-ROC (in an one-vs-all capacity). Each target was evaluated separately.

### Hyperparameter Optimization, Model Fine-Tuning, and Model Selection

For hyperparameter optimization, we performed an iterative gridsearch across two learning rates, three batch sizes, and three random seeds (used for train/validation split). Due to memory limitations, the maximum number of input tokens per document that we were able to implement was 2,048 input tokens. We used 512 input tokens for CB (the maximum allowed by the CB model), but for CBB we experimented with 512, 1,024, and 2,048 (the maximum allowed by our hardware). We fine-tuned each model for 30 epochs. Run-time of CBB experiments was substantially longer than that of CB experiments, with 2,048 input token CBB (CBB-2048) instantiations taking almost 24 hours of training run-time per parameter combination.

We evaluated model performance depending on TCGA validation set AU-ROC, selecting the best final model per target based on this metric. We found that CBB-2048 was the best model type for T14 and N03 targets, whereas CBB-1024 was the best for the M01 target (Table S2). The final TNM models are made publicly available through Huggingface [23], which is a widely used Python library for publishing and downloading large language models.

### Characterization of CUIMC Pathology Report Dataset

We retrieved all reports from the CUIMC pathology report database, between 2010-2019. We removed empty reports and “outside consultation” reports. We selected for reports with the “surgical pathology” label, as this label indicated histopathology slide analysis in contrast to other report types generated by the pathology department. Report text remained intact, not pre-processed. TNM stage annotation data were located in a separate metadata table, derived from the tumor registry. We selected for patients with non-empty TNM values.

We employed three attributes to match report text to patient TNM annotation: patient ID, report date (matched to TNM diagnosis date), and TNM-primary site (Figure S4). Patient ID was matched exactly across the two databases. For date-matching, we allowed up to 90 days between report-date and diagnosis-date, as there is a lead-time/delay between pathologist documentation and official tumor registry stage extraction. We observed that the number of reports overall, as well as the number of reports per patient, increased as the time-window was expanded from 0 to 90 days. Additionally, we observed that a single patient may have multiple pathology reports potentially associated with a given TNM annotation, within the same time-window. We therefore imposed an additional matching requirement to ensure report-annotation relevancy, selecting the most “relevant” report as that which has the greatest number of report string matches to the TNM-associated “primary site” value. At this stage, the vast majority of patients were associated with a single TNM-report match. However, in the event that multiple reports were equally relevant, we concatenated reports to ensure that all relevant TNM-information would be captured.

In the final CUIMC dataset, most reports had associated T14 annotation, and the least number of reports had M annotation, similar to the TCGA dataset (Table S3A). We tabulated the class-imbalance for each target (Table S3C). We found that T4 and N3 are the least-prevalent classes per target, as was the case for the TCGA report set (Figure 1C). We also found that the proportion of M1 is higher in the CUIMC dataset (20.1%) as compared to the TCGA dataset (6.7%). The range of diseases is larger for the CUIMC reports as compared to TCGA reports: The TCGA dataset ranged from 21-23 cancer types, whereas the CUIMC dataset spans 40-42 primary sites (although these terms are not directly comparable). We plotted the primary site distribution for each target report set (Figure S4), finding that the distributions are similar across the three targets. As in the TCGA dataset, “breast” and “lung” are two of the most prevalent cancer sites, across all three targets. Finally, using the CBB tokenizer, we computed token statistics for each target-dataset (Table S3B). Overall, we found that CUIMC pathology reports were longer than TCGA pathology reports, both on average and at maximum report length.

### Application of TCGA-Trained Models to CUIMC Dataset

TNM models were applied directly to the entire CUIMC report set (without any additional fine-tuning). As before, we calculated AU-ROC to evaluate model performance. We found that, as for TCGA validation and held-out test sets, M01 was the least well-performing model (as compared to the T14 and N03 models).

We compared the CUIMC performance of our TNM models to those of Abedian et al. (2021) [5], which was the most comparable to ours in terms of the use of pathology report text as sole input, the predicted TNM target value ranges (T14, N03, and M01), and the inclusion of multiple cancer types in both train and test sets. Abedian et al. reported F1, rather than AU-ROC. We computed F1 for our models and compared our results to the pan-cancer test set results in [5] (Table S3D). We found that our T14 model performed on-par with [5], our N03 model performed somewhat better, and our M01 model performed substantially better than the equivalent model in [5].

We performed three additional experiments to probe our external validation results. First, although we found that the CBB model-type achieved the best performance on the TCGA report set, we were interested in whether this result would hold for CUIMC reports. To test this, we applied the best-performing TCGA-trained CB model to the CUIMC report set to predict the N03 target. There was a large difference in performance across all evaluation metrics, including overall macro and per-class AU-ROC between CB and CBB (Table S4). CBB likely performed better than CB due to its increased complexity as well as its increased input token size (Table S3B).

Second, we tested whether our primary parameter for report-diagnosis matching, number of days between diagnosis and report date, had any impact on CUIMC performance. We ran the TCGA-trained models on CUIMC data for each target separately for 0, 10, and 30 days; we compared the results to the performance we achieved with 90 day report-matching (Figure S5). In this sensitivity analysis, we found that AU-ROC remained stable as the number of days was varied. For the multi-class targets, T14 and N03, we plotted per-class changes over time, finding that there is a slight increase in per-class AU-ROC as the number of days increases. The magnitude of AU-ROC increase across number of days varies by class. The least prevalent classes (e.g., T4 and N3) have the largest gain in AU-ROC as number of days increases; this is likely due to the increased likelihood of report relevance as number of days increases.

Finally, we tested the removal of patient-identifying information (PHI), such as medical record number, date of birth, etc., from the preamble of each report for the T14 target. In the CUIMC dataset, most of the patient-identifying text was located in the first few lines of each report (whereas diagnosis information was not typically contained in this preamble section). Our hypothesis was that the model may perform better without extraneous patient details, particularly as these types of details had not been seen by the model when trained on the de-identified TCGA report set. However, we observed only a 0.0001 AU-ROC gain when PHI was removed (Table S5). We determined that PHI removal was not necessary for external validation, as increased pre-processing effort would potentially lead to only a very small performance gain.

## Data Availability

Python scripts used in this study can be found on github. Models generated by this study can be found on Hugging Face. TCGA pathology report text can be found on github. CUIMC pathology reports are not available due to HIPAA compliance.

https://github.com/tatonetti-lab/tnm-stage-classifier

https://github.com/tatonetti-lab/tcga-path-reports

https://huggingface.co/jkefeli/CancerStage_Classifier_T

https://huggingface.co/jkefeli/CancerStage_Classifier_N

https://huggingface.co/jkefeli/CancerStage_Classifier_M

## Acknowledgements

We would like to acknowledge the help of Benjamin May for providing pathology reports from the CUIMC data warehouse. JK and NPT are supported by NIH NIGMS R35GM131905.

## Code Availability

Python scripts used in this study can be found on Github:

https://github.com/tatonetti-lab/tnm-stage-classifier

Models generated by this study can be found on Huggingface:

https://huggingface.co/jkefeli/CancerStage_Classifier_T

https://huggingface.co/jkefeli/CancerStage_Classifier_N

https://huggingface.co/jkefeli/CancerStage_Classifier_M

## Author Contributions

JK conceived the study, performed data analysis, code, manuscript. NT performed study supervision and edited manuscript.

## Data Availability

TCGA pathology report text can be found at https://github.com/tatonetti-lab/tcga-path-reports CUIMC pathology report text is not available due to HIPAA/PHI.

## Supplementary Material

**Figure S1.**
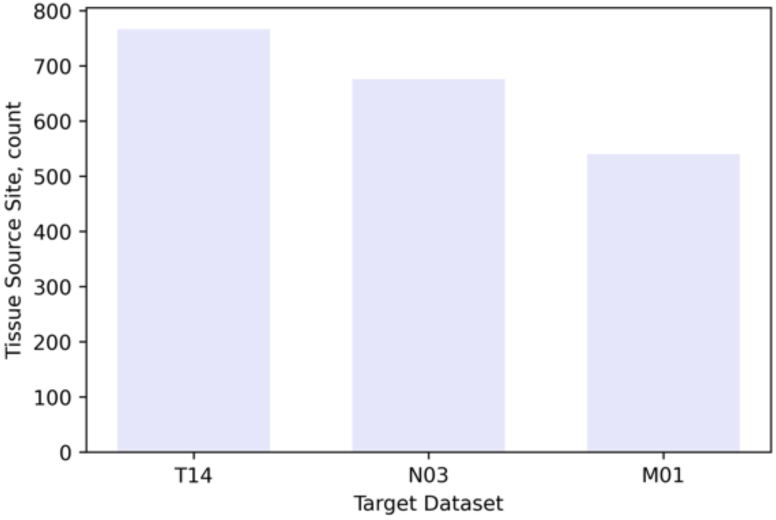
Distribution of number of tissue source sites across TCGA for each target dataset. Each tissue source site represents a different institution that contributed both slides and pathology reports to the TCGA dataset. Within each institution, there are a variety of pathologists contributing reports. As a result, each target dataset contains reports with a wide range of formatting, writing styles, and abbreviations.

**Table S1.** Tokenization statistics for TCGA training set reports. The CB Tokenizer was used. Included the number (n) and proportion (p) of reports above each threshold token limit. Model CB has a per-report 512 input token limit, whereas model CBB has an input threshold of 4,096 tokens per report.

| Maximum Tokens Threshold | Reports above Threshold (n) | Reports above Threshold (p) |
| --- | --- | --- |
| 512 | 5345 | 66.0% |
| 1024 | 3122 | 38.6% |
| 2048 | 1045 | 12.9% |
| 4096 | 55 | 0.7% |

**Table S2.**
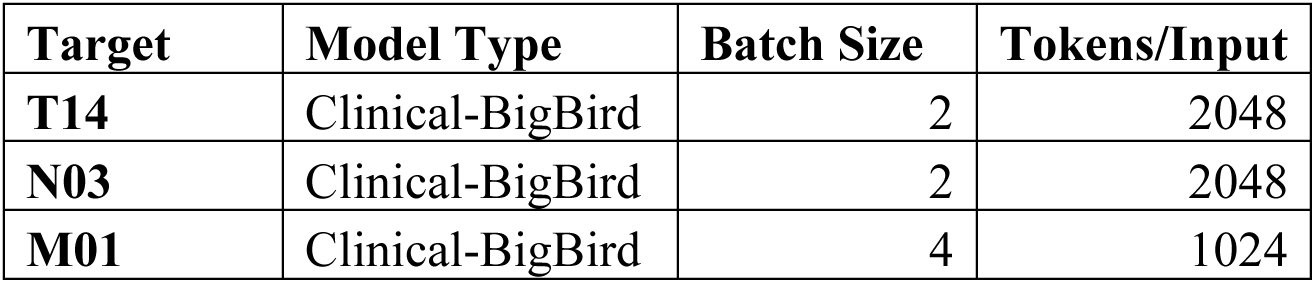
Best-performing model parameters, by classification target, based on TCGA validation set performance. Included parameters are model type, batch size, and maximum tokens per input report. Batch size was limited by memory constraints. ClinicalBERT was not found to be best-performing for any of the target classification tasks.

**Figure S2.**
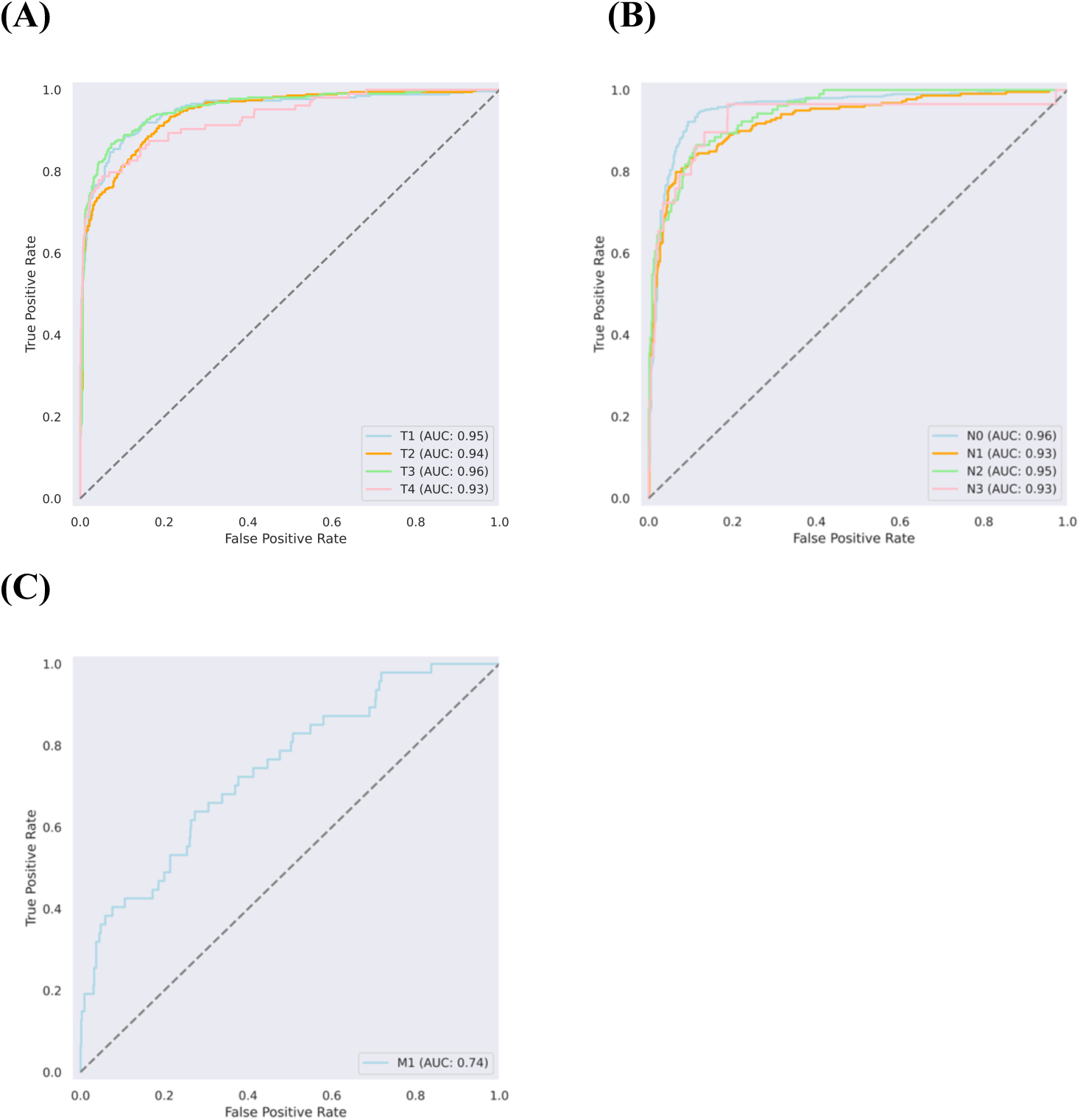
Overall model performance on held-out TCGA test set. ROC curves for (A) T14, (B) N03, and (C) M01 models. Individual AU-ROC is captured in each plot legend. For (A) and (B), each curve corresponds to per-class performance, calculated as one-versus-all.

**Figure S3.**
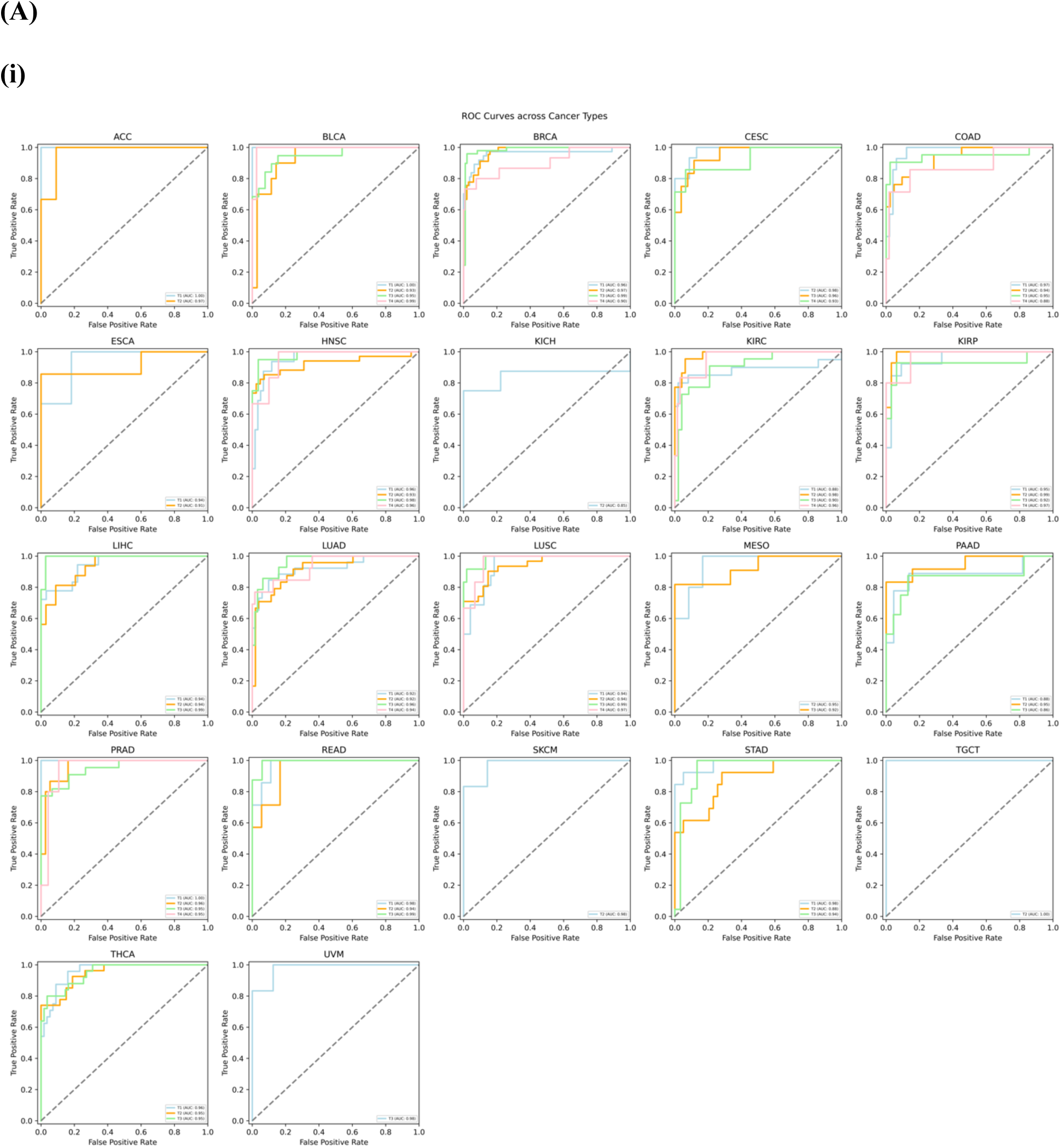
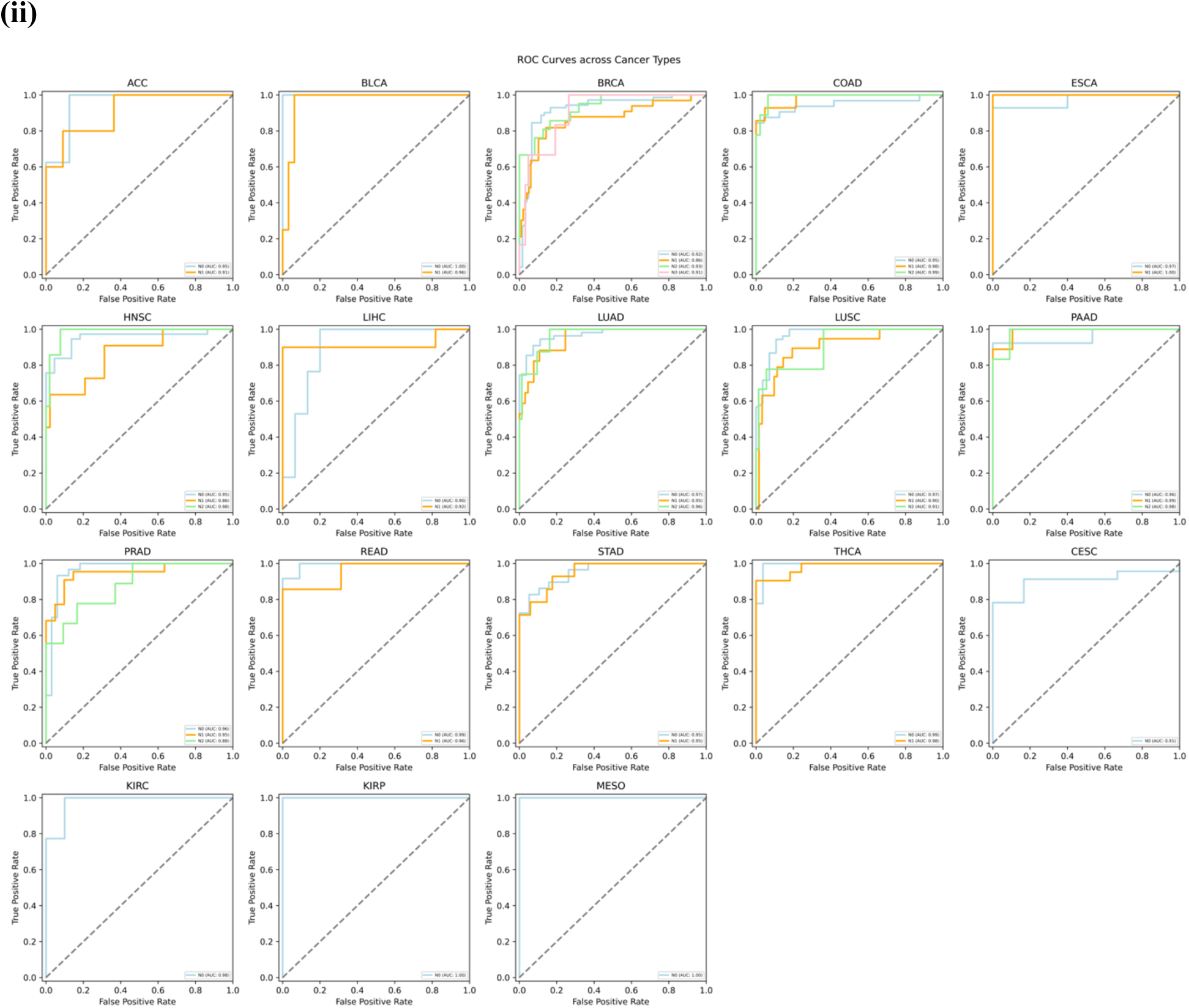
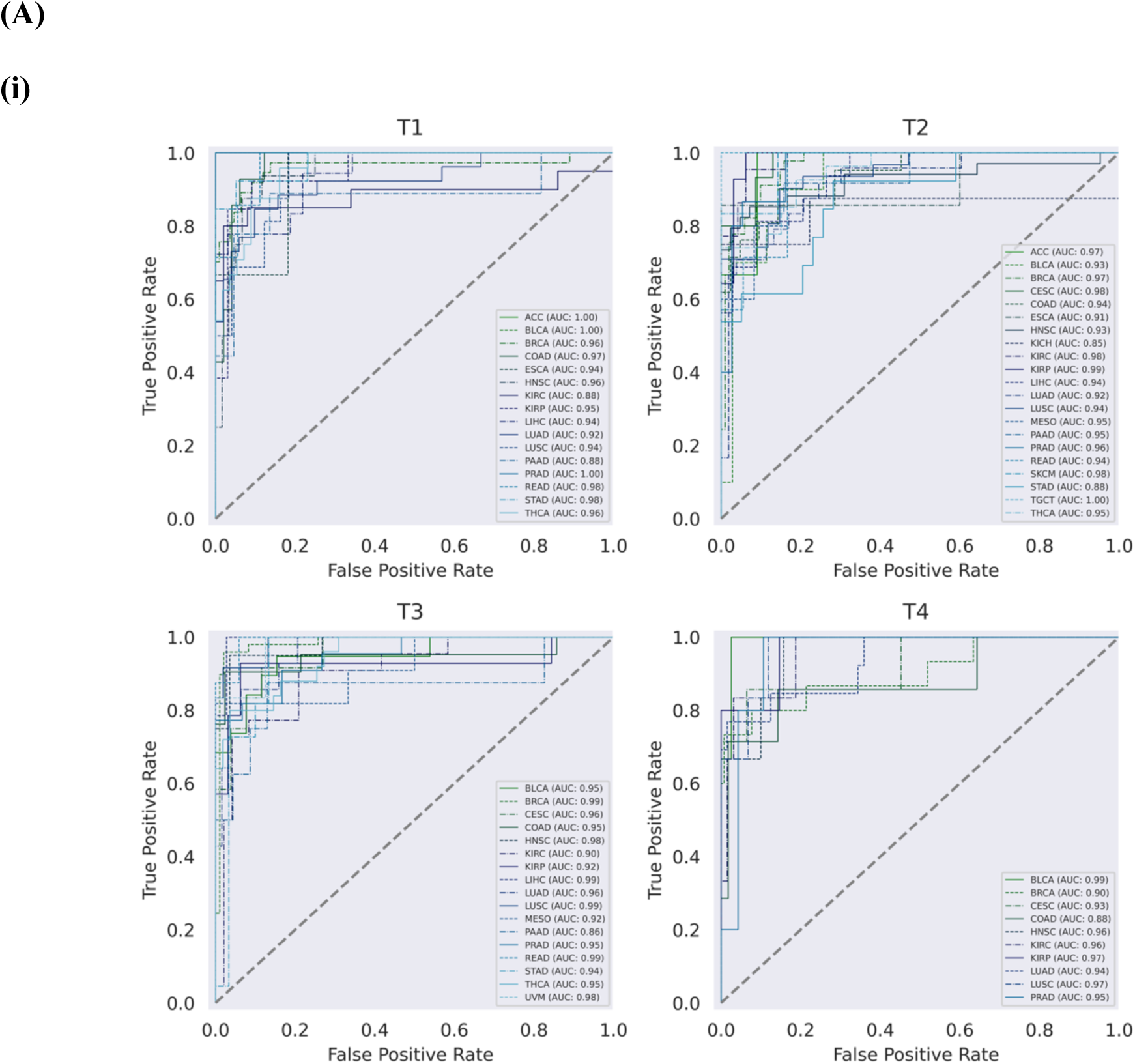
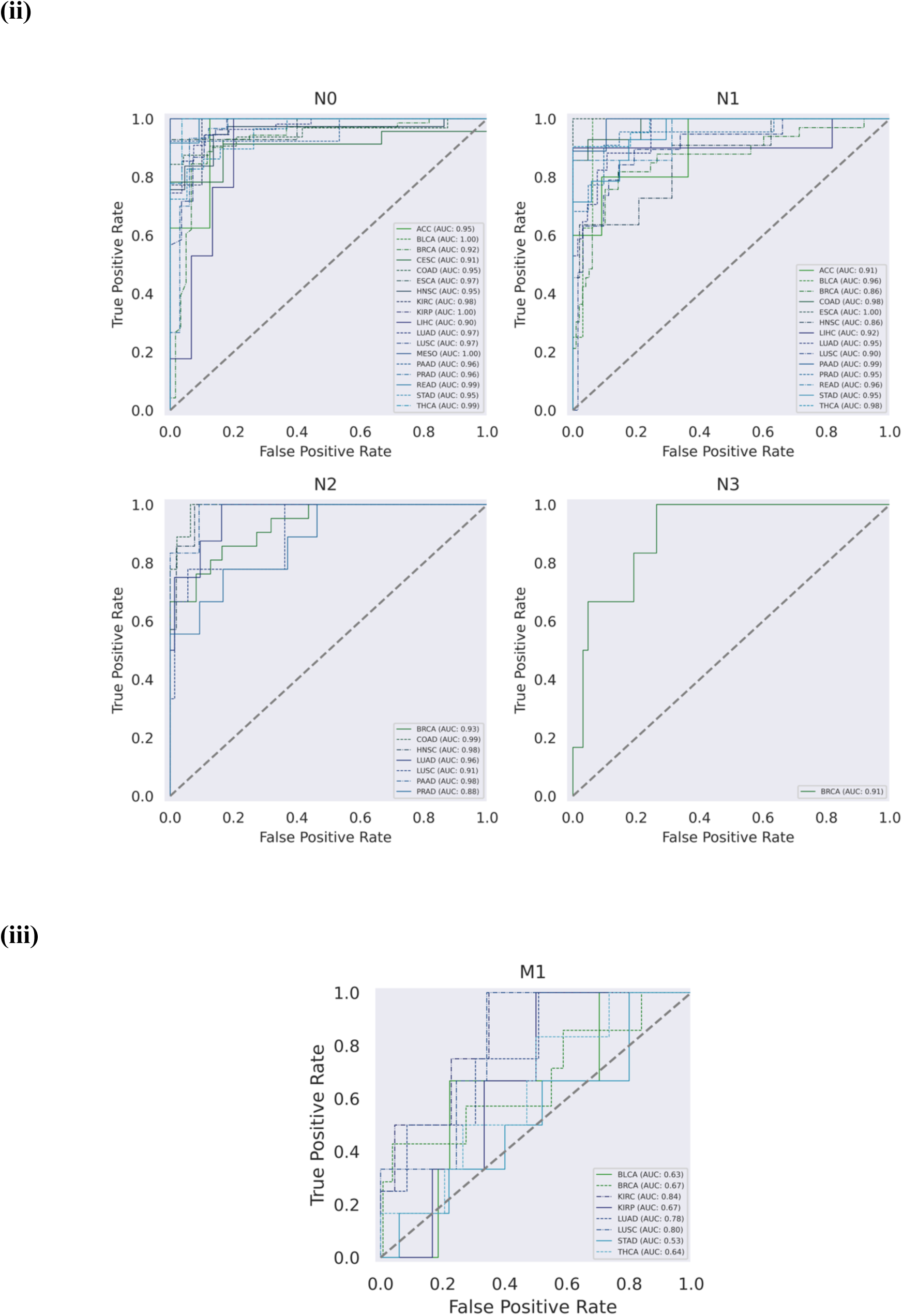
Per-class and per-cancer type best-model performance on held-out TCGA test set. (A) Per-cancer type, across classes. (i-ii) T14, N03. (B) Per-class, across cancer types. (i-iii) T14, N03, M01. At least 5 examples per class in the test set were required for cancer type inclusion, except for M01, for which the threshold was reduced to 3 test examples per class due to low-data availability. AU-ROC is presented in each plot legend.

**Table S3.**
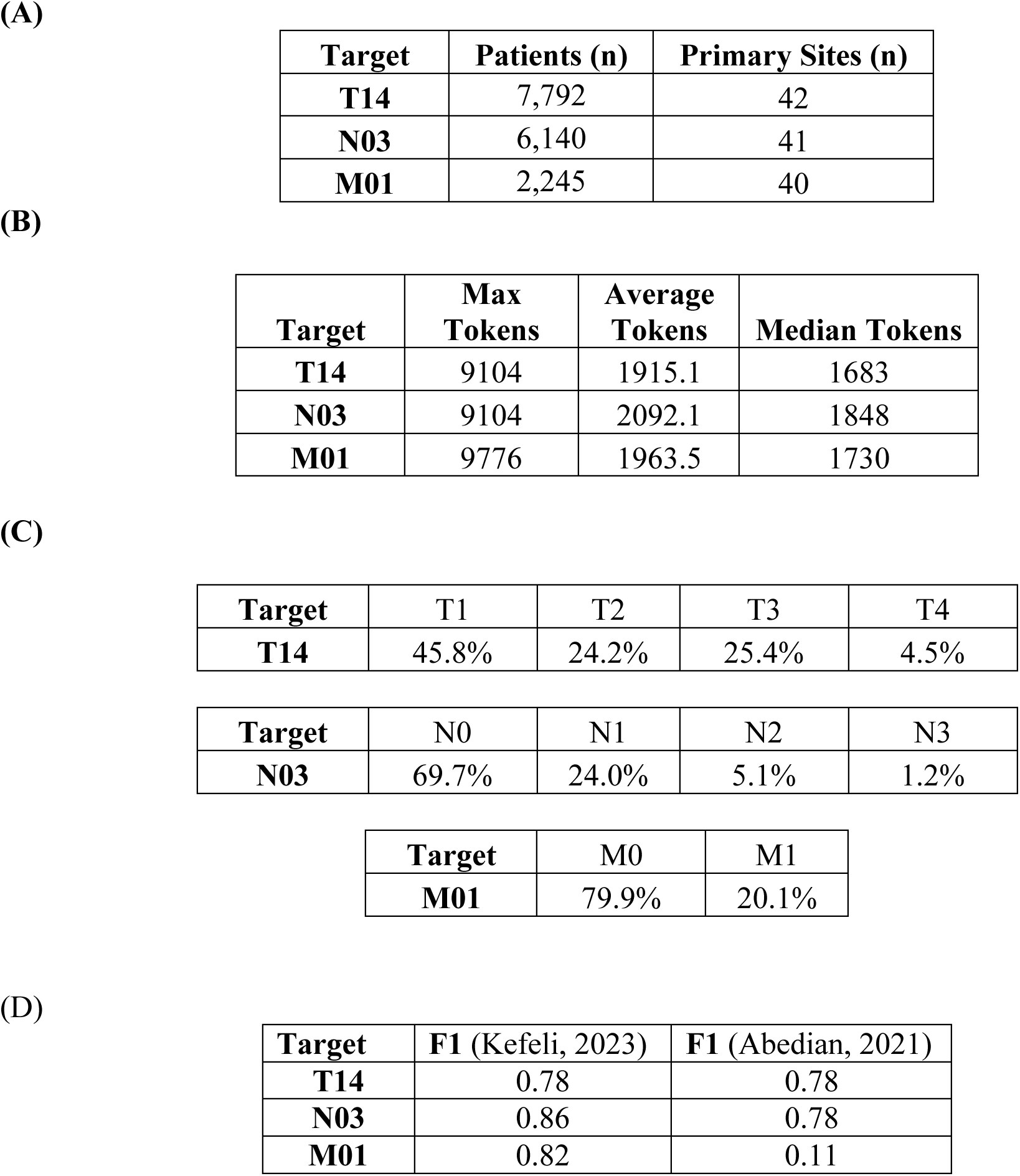
CUIMC report set characterization. (A) Compiled CUIMC report statistics, number of patients and primary cancer sites. (B) Tokens per input for CUIMC pathology reports, using CBB Tokenizer. (C) CUIMC class distribution, by target (T, N, M). (D) Performance of TNM models on CUIMC data, as compared to TNM model performance in (Abedian et al., 2021), across all cancer types. For (Abedian et al., 2021) results, we selected the pan-cancer “random subtypes” test set; this matched most closely with the inclusion of all subtypes in the CUIMC dataset. F1 is micro-computed.

**Figure S4.**
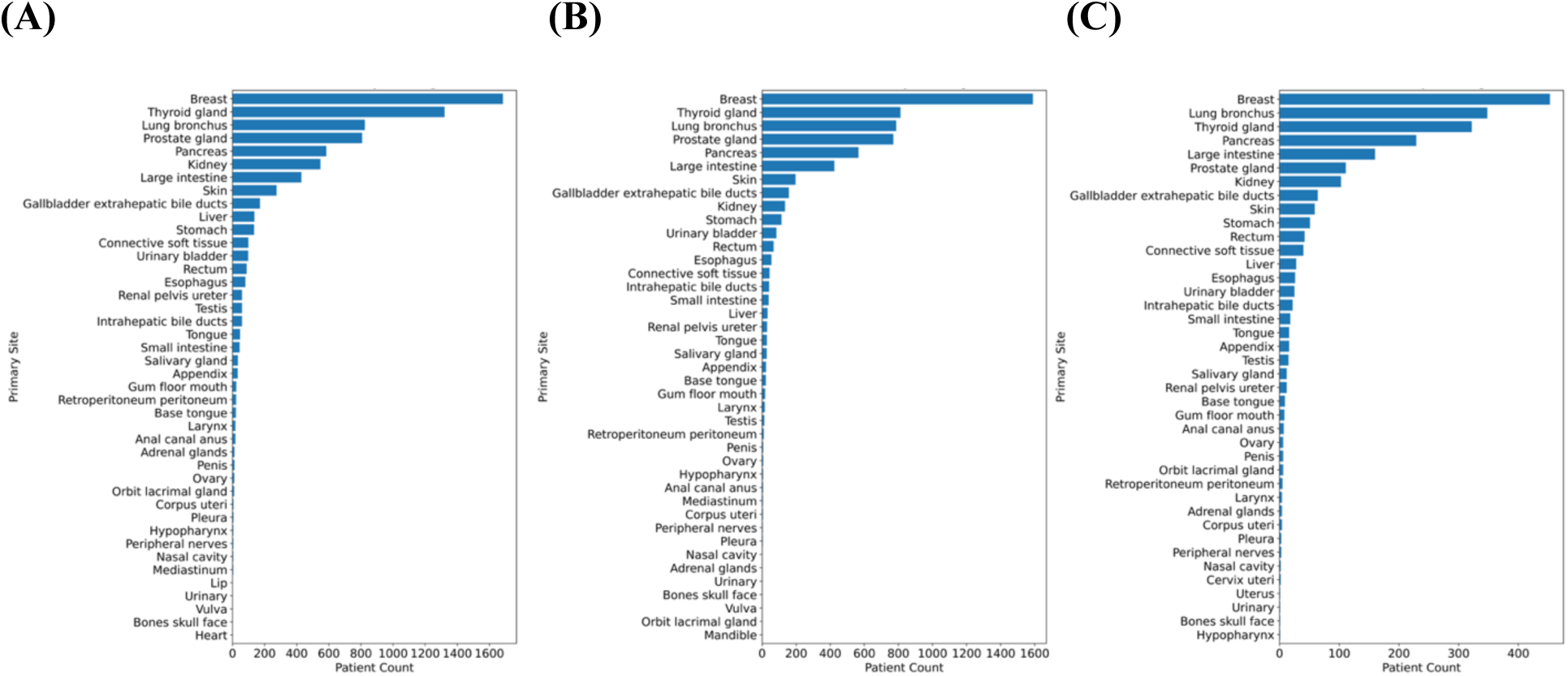
Primary site distribution of CUIMC reports. (A) T14, (B) N03, (C) M01.

**Table S4.** CUIMC Performance on N03 target with TCGA-trained ClinicalBERT, 512 input tokens (CB-512), and TCGA-trained Clinical-BigBird, 2,048 input tokens (CBB-2048). AU-ROC macro-computed.

| Model | AU-ROC | F1 Micro | AU-ROC (N0) | AU-ROC (N1) | AU-ROC (N2) | AU-ROC (N3) |
| --- | --- | --- | --- | --- | --- | --- |
| CB-512 | 0.779 | 0.688 | 0.782 | 0.782 | 0.764 | 0.789 |
| CBB-2048 | 0.912 | 0.863 | 0.937 | 0.922 | 0.916 | 0.872 |

**Table S5.** CUIMC Performance on T14 target, with and without PHI preamble per report. AU-ROC macro-computed.

| Condition | AU-ROC | F1 Micro |
| --- | --- | --- |
| With PHI | 0.9415 | 0.7767 |
| Without PHI | 0.9416 | 0.7862 |

**Figure S5.**
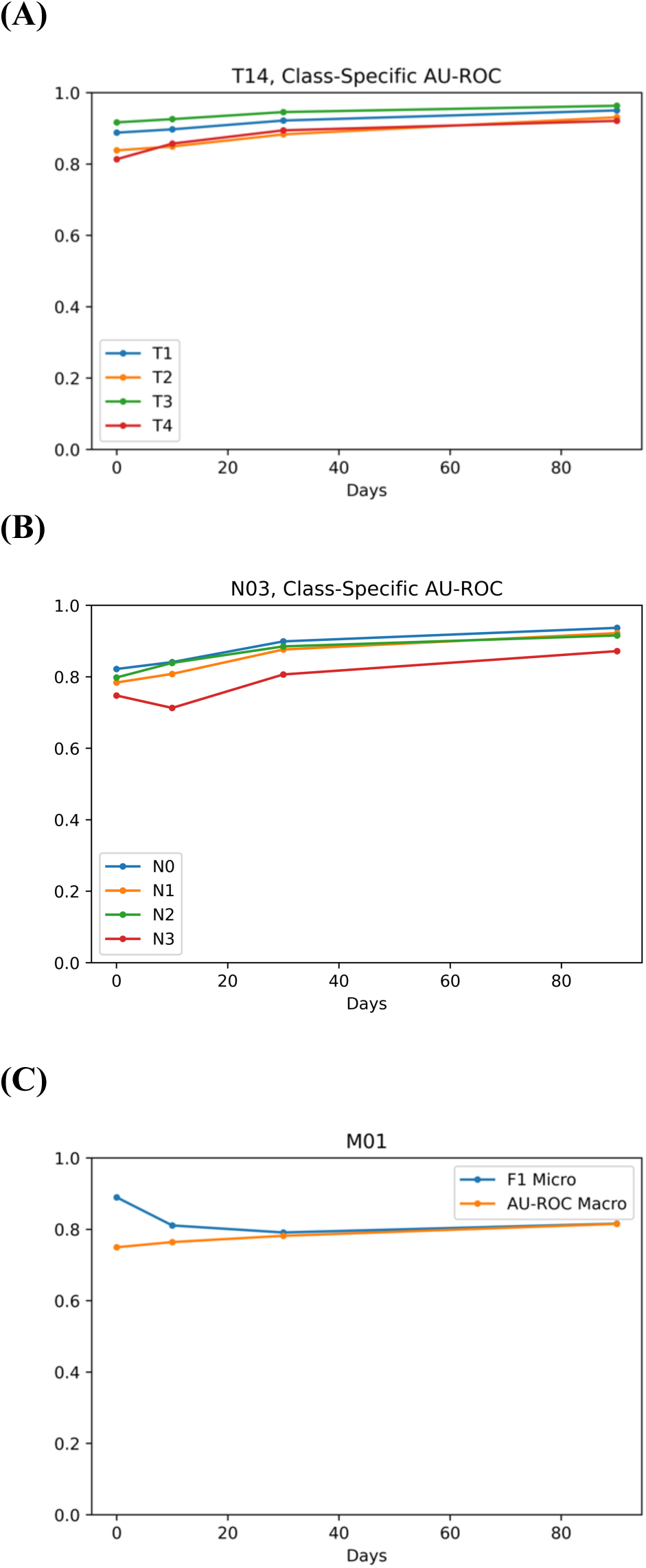
Sensitivity analysis, AU-ROC values with increasing number of days (time-windows) between pathology report and documented diagnosis (for report-matching process, see Methods). (A) T14, (B) N03, (C) M01. For (A) and (B), AU-ROC is computed per-class for (C), which is binary, AU-ROC and F1-micro are plotted.

